# Statistics based predictions of coronavirus 2019-nCoV spreading in mainland China

**DOI:** 10.1101/2020.02.12.20021931

**Authors:** Igor Nesteruk

**Affiliations:** Institute of Hydromechanics, National Academy of Sciences of Ukraine, Zheliabova 8/4, UA-03680 Kyiv, Ukraine; National Technical University of Ukraine “Igor Sikorsky Kyiv Polytechnic Institute”, Prosp.Peremohy 37, UA-03056, Kyiv, Ukraine

**Keywords:** coronavirus epidemic in China, coronavirus 2019-nCoV, mathematical modeling of infection diseases, SIR-model, parameter identification, statistical methods

## Abstract

**Background:** The epidemic outbreak cased by coronavirus 2019-nCoV is of great interest to researches because of the high rate of spread of the infection and the significant number of fatalities. A detailed scientific analysis of the phenomenon is yet to come, but the public is already interested in the questions of the duration of the epidemic, the expected number of patients and deaths. For long time predictions, the complicated mathematical models are necessary which need many efforts for unknown parameters identification and calculations. In this article, some preliminary estimates will be presented.

**Objective:** Since the reliable long time data are available only for mainland China, we will try to predict the epidemic characteristics only in this area. We will estimate some of the epidemic characteristics and present the most reliable dependences for victim numbers, infected and removed persons versus time.

**Methods:** In this study we use the known SIR model for the dynamics of an epidemic, the known exact solution of the linear equations and statistical approach developed before for investigation of the children disease, which occurred in Chernivtsi (Ukraine) in 1988-1989.

**Results:** The optimal values of the SIR model parameters were identified with the use of statistical approach. The numbers of infected, susceptible and removed persons versus time were predicted.

**Conclusions:** Simple mathematical model was used to predict the characteristics of the epidemic caused by coronavirus 2019-nCoV in mainland China. The further research should focus on updating the predictions with the use of fresh data and using more complicated mathematical models.

## INTRODUCTION

Here, we consider the development of epidemic outbreak cased by coronavirus 2019-nCoV (see e.g., [1-3]). Since the reliable long time data are available only for mainland China, we will try to predict the number of victims *V* of this virus only in this area. The first estimations of *V* exponential growth, typical for the initial stages of every epidemic (see e.g., [4]) have been done in [3]. For long time predictions, more complicated mathematical models are necessary. For example, a susceptible-exposed-infectious-recovered (SEIR) model was used in [2]. Nevertheless, the complicated models need more efforts for unknown parameters identification. This procedure may be especially difficult, if reliable data are limited.

In this study, we use the known SIR model for the dynamics of an epidemic [4-8] To the parameter identification, we will use the exact solution of the SIR set of linear equations and statistical approach developed in [4]. These methods were applied for investigation of the children disease, which occurred in Chernivtsi (Ukraine) in 1988-1989. We will estimate some of the epidemic characteristics and present the most reliable dependences for victim numbers, infected and removed persons versus time.

## MATERIALS AND METHODS

### Data

We shall analyze the daily data for the number of confirmed cases in mainland China, which origins from China National Health Commission [1]. We show in the table Table 1 the corresponding time moments *t*_*j*_ from 0 to 24 and the number of victims *V*_*j*_ (confirmed cumulative cases of coronavirus 2019-nCoV infection), which were used for calculations. Table 1 shows that the precise time of the epidemic beginning *t*_0_ is unknown. Therefore, the optimization procedures have to determine the optimal value of this parameter as well as for other parameters of SIR model.

**Table 1.**
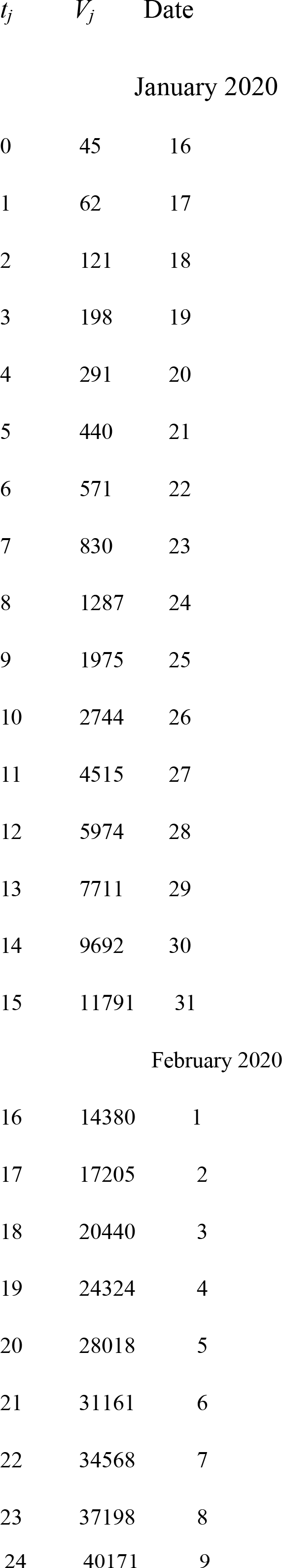
The information from official table of from China National Health Commission [1]. The corresponding time moments *t*_*j*_ and the number of victims V_j_ (confirmed cumulative cases of coronavirus 2019-nCoV infection), which were used for calculations.

### Exact solution of SIR-equations

The SIR-model for an infectious disease can be written as follows, [6,7]:

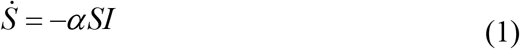

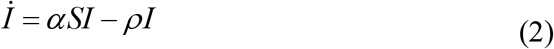

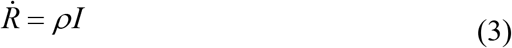

The number of susceptible persons is *S*, infected - *I*, removed -*R*; the infection and immunization rates are *α* and *ρ* respectively. Since 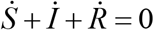 (see, Eqs. (1-3)), the sum *N* = *S* + *I* + *R* must be constant for all moments of time and can be treated as the amount on susceptible persons before the outbreak of an epidemic, since *I* = *R* = 0 at *t* < *t*_0_. It must be noted that the constant *N* is not the volume of population *N*_*total*_, but only the initial number if people sensitive and not protected to some specific disease. In particular, the ratio *N* / *N*_*total*_ may be rather small. For example, the number of people on the board of Diamond Princess is 3711, and the number of confirmed cases is 70 (February 10, 20200). It means that the percentage of susceptible can be estimated by 1.89%.

To determine the initial conditions for the set of equations (1-3), let us suppose that

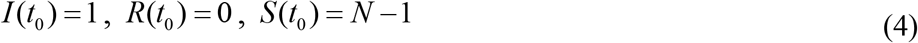

It follows from (1) and (2) that

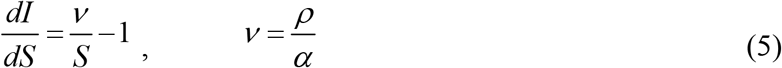

Integration of (5) with the initial conditions (4) yields:

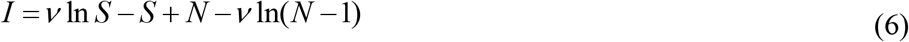

Function *I* has a maximum at *S* =*ν* and tends to zero at intinity, see [6, 7]. In comparison, the number of susceptible persons at infinity *S*_∞_ > 0, and can be calculated with the use of (6) from a non-linear equation

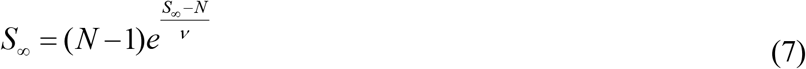

In [4] the equations (1-3) were solved by introducing the function *V* (*t*) = *I* (*t*) + *R*(*t*), corresponding to the number of victims. The integration of corresponding equation:

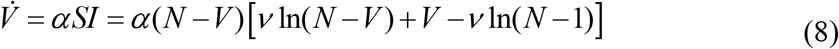

yields:

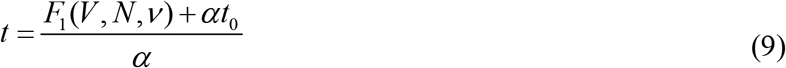

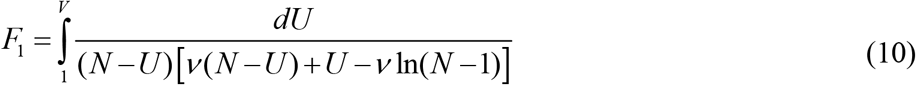

Thus, for every set of parameters *N, ν, α, t*_0_ and a fixed value of *V* the integral (10) can be calculated and the corresponding moment of time can be determined from (9). Then *I* can be calculated from (6) by putting *S=N-V* and function *R* from *R=V-I*.

### Statistical approach for parameter identification. Linear regression

As in paper [4], we shall use the fact that the random function *F*_1_ (*V, N,ν*) has a linear distribution with (see (9)). Then we can apply the linear regression (see [9]) for every pair of parameters *N* and *ν* and the corresponding values of *t*_0_ and *α*. The optimal (the most reliable) values of *N* and *ν* correspond to the maximum value of the correlation coefficient *r* (see [4]).

## RESULTS

The optimal values of parameters were calculated:

*N*=90611; *ν* =65546.5; *α* =1.477985357571669e-05; *t*_0_ = -7.720998173432072.

The corresponding correlation coefficient is very high *r*=0.997966487046645. The solution of (7) yields the value *S*_∞_ = 45579. The corresponding number of infected *I*, susceptible *S* and removed *R* persons versus time (starting from January 16, 2020) were calculated and shown in Fig. 1. The blue line represent the number of victims *V=I*+*R* and is in good agreement with confirmed number of victims *V=I*+*R* (blue markers, see [1]).

**Fig. 1.**
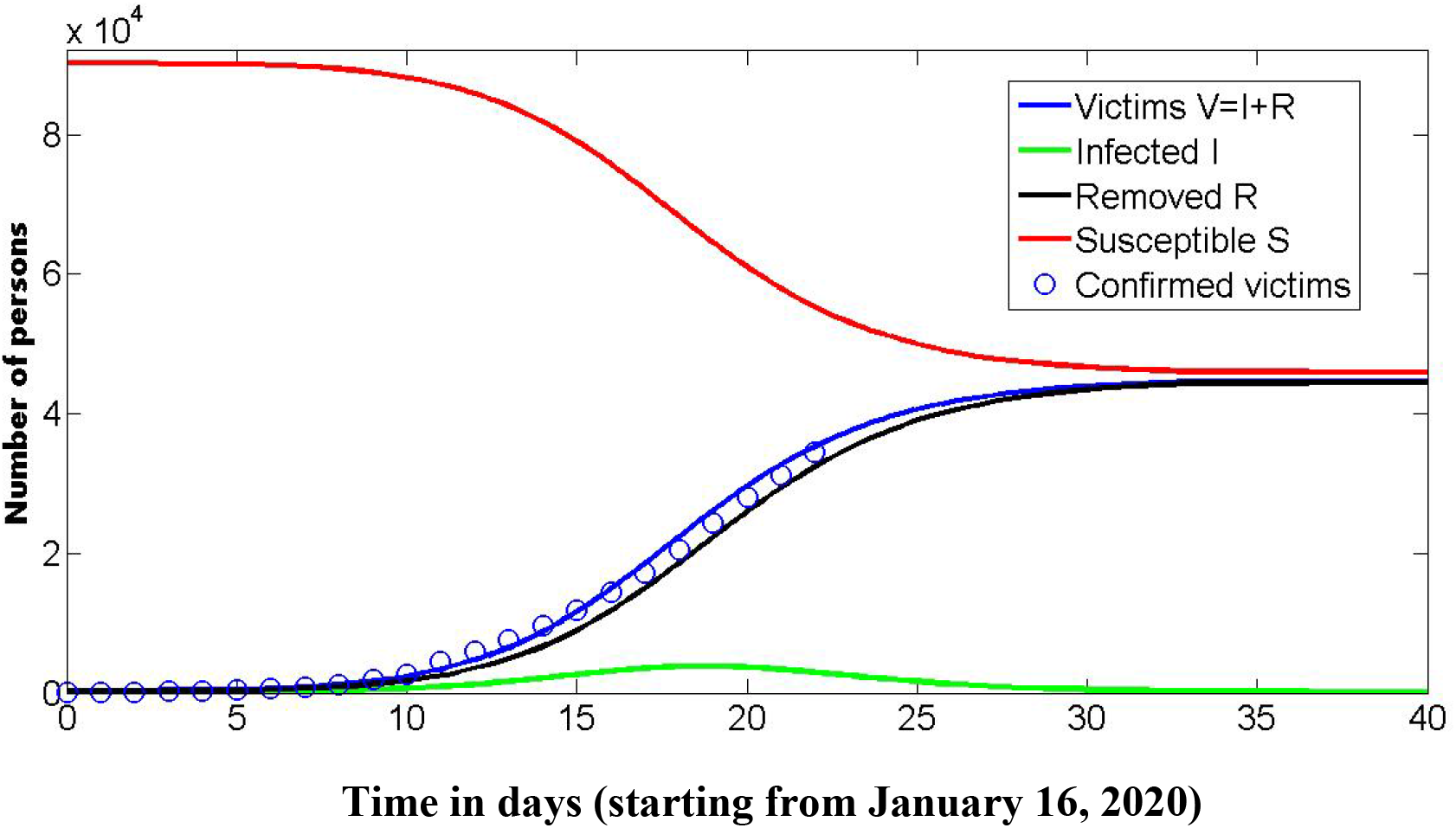
Numbers of infected *I* (green line), susceptible *S* (red line) and removed *R* (black line) persons versus time in days (starting from January 16, 2020). The blue line represents the number of victims *V=I*+*R*. Blue markers show the confirmed number of victims, reported by China National Health Commission [1].

## DISCUSSION

The obtained value of the correlation coefficient if very close to unit 1. This fact can make the results rather reliable. Nevertheless, this the value of *r* is very close to the optimal one *r*=0.997966487046645 for the values of parameters located in the vicinity of the optimal point *N*=90611; *ν* =65546.5, since the maximum of regression coefficient at this point is not sharp. This fact can question the procedure of the parameter identification. The calculations must be refreshed after obtaining new data.

Another weak feature of the method is connected with the fact that the estimation of susceptible persons, who are still present in the population *S*_∞_ = *N* −*V*_∞_ =45579, is very large. It means that these persons can couch the infection. Such situation needs additional analysis, in particular, with the use of more complicated models (see, e.g., [10]).

## CONCLUSIONS

Simple mathematical model was used to predict the characteristics of the epidemic caused by coronavirus 2019-nCoV in mainland China. The further research should focus on updating the predictions with the use of fresh data and using more complicated mathematical models.

## Data Availability

The data used are available in the text

## Acknowledgements

I would like to express my sincere thanks to professors Dirk Langemann (Techniche Universitaet Braunschweig) and Juergen Prestin (Universitaet zu Luebeck) for their support in developing the used optimization approach. I would like to thank also professors Alberto Redaelli, Giuseppe Passoni and Gianfranco Fiore (Politecnico di Milano), S. Pereverzyev (RICAM, Linz, Ausria) for involving me in very interesting biomedical investigations in frames of EU-financed Horizon-2020 projects EUMLS (Grant agreement PIRSES-GA-2011-295164-EUMLS) and AMMODIT (Grant Number MSCA-RISE 645672).

